# Crowd signals: Early detection of disease outbreaks using real-time healthcare occupancy data

**DOI:** 10.1101/2025.09.10.25335525

**Authors:** Jose Deney Araújo, Juan Carlo Santos e Silva, Marcelo A S Bragatte, Erick Rodrigues Sousa, Isaac N Schrarstzhaupt, Ester C. Sabino, Helder I Nakaya, Carolina dos Santos Lázari, João Renato Rebello Pinho, Gerson Oliveira Penna, Jorge Kalil, Mariangela Simão, Anderson Fernandes Brito, Vanderson Sampaio

## Abstract

Early detection of disease outbreaks is critical for effective public health response, yet traditional surveillance systems often suffer from delayed reporting. Here, we investigate whether real-time occupancy data from healthcare facilities can act as an early warning indicator of possible outbreak activity. We analyzed occupancy trends from 17 emergency care units in the São Paulo metropolitan area and compared them with national surveillance data for infectious diseases, including SARS-CoV-2 and dengue virus. Dynamic time warping and Granger causality tests demonstrated that occupancy patterns anticipate infection dynamics with a mean lead time of three weeks. Early warning signals of three epidemiological events were identified as deviations from average occupancy. Local indicators of spatial association revealed persistent overcrowding hotspots in later outbreak stages, highlighting regions where sustained healthcare monitoring and surveillance remain necessary. These findings demonstrate the potential of privacy-safe passive occupancy data to support timely epidemic surveillance.

## Introduction

Global health crises trigger widespread social, economic, and political disruptions, with healthcare facility overcrowding standing as a key pressure point for system resilience ^1^. Beyond the direct burden on care delivery, overcrowding is associated with operational inefficiencies, negative patient outcomes, prolonged wait times, and reduced bed availability ^2,3^. Environmental hazards such as heatwaves, wildfires, and floods have been recognized as important sources of pressure on health systems. These non-infectious drivers of overcrowding highlight the need for sensitive surveillance approaches that can monitor fluctuations in healthcare facility capacity ^4–6^. Recently, electronic health records were used to estimate facility occupancy, but limitations in accessibility and the interval between data collection and analysis constrain their utility for rapid epidemiological response ^7^. While pathogen-specific data remains essential for diagnosis and response, planning requires time ^8^. From an operational perspective, sensitive early signals of anomalous healthcare usage can provide timely insights to support rapid decision-making and resource allocation ^9^. Therefore, the ability to detect early warnings of disease outbreaks based on healthcare unit occupancy represents a potential innovation that could add a new layer to epidemic intelligence and significantly enhance public health surveillance ^10^.

Health systems worldwide use multiple surveillance strategies to track infectious diseases. These include passive notification systems for notifiable diseases such as COVID-19, dengue, and yellow fever ^11^; sentinel surveillance from selected healthcare facilities, syndromic and laboratory-based surveillance ^12^. However, while these methods are well-established, they often suffer from limitations such as delayed reporting, high infrastructure demands, and lack of real-time availability ^13^. Currently, many digital surveillance tools incorporate artificial intelligence to mine open-source data from social media or news, yet these approaches can suffer from noise and limited clinical relevance ^14^. Despite advances, most official notification systems still require several days to identify and report outbreaks, causing delays that can undermine timely interventions ^15^. While surveillance systems exist to track infectious diseases, few tools effectively monitor real-time stress on healthcare infrastructure. In this context, there is a pressing need for complementary systems that harness alternative data streams to enhance the speed, sensitivity, and scalability of epidemic detection.

In this study, we present a novel method that leverages real-time occupancy data from healthcare facilities, made available by Google Maps^TM^, to detect early warning signals of potential outbreak activity. This data, derived from anonymized and aggregated user location metrics, provides a dynamic proxy for occupancy levels in clinical settings. Sudden increases in facility occupancy were associated with intensified pathogen circulation, making this a powerful source for early outbreak detection. We tested this hypothesis by analyzing occupancy patterns from 17 public urgent care units in the São Paulo metropolitan area. By applying Dynamic Time Warping (DTW) and Granger Causality analyses, we demonstrated that occupancy trends are associated with confirmed cases of respiratory pathogens and dengue virus (DENV) infections by up to five weeks. Unsupervised spatial analysis uncovered persistent high levels of healthcare unit occupancy in the later stages of the epidemiological event, pointing to areas that continued to experience system strain and required ongoing monitoring. Our findings show that the use of privacy-safe passive mobility data can offer a timely, low-cost, and scalable solution to enhance epidemic surveillance and public health responses.

## Results

### Healthcare occupancy patterns reflect epidemiological events timing

Across seven cities in the São Paulo metropolitan area, aggregated occupancy data from 17 healthcare units reflected population-level dynamics over the period from July 15, 2023, to October 12, 2024 (Figure 1A and Supplementary Data 1). Temporal relationships between weekly occupancy averages and laboratory-based surveillance data (Figure 1B) revealed whether historical trends in one time series exhibited lagged associations with the other, using time-lagged modeling approaches that account for directional dependencies (Figure 1C). Laboratory surveillance data included SARS-CoV-2, DENV, and other respiratory viruses of public health importance (RV: SARS-CoV-2, Influenza virus A/B, and respiratory syncytial virus).

Temporal distribution of occupancy levels revealed notably coherent dynamics across the healthcare units, despite their independent selection and heterogeneous contexts (Figure 2A). Three distinct periods concentrated the highest occupancy levels across units: from July to October 2023, from December 2023 to May 2024, and from August to September 2024. The occupancy average trajectory highlights the collective shifts and further supports the similar patterns among the units (Figure 2A, lower panel). These periods of elevated occupancy partially overlap with potential outbreak intervals (Figure 2B-C), suggesting consistent temporal patterns in occupancy surges that may reflect broader epidemiological dynamics. Despite using occupancy averages, waves of overcrowding were consistently observed across the 17 monitored units, underscoring the synchronized signal that can be used to track health system pressures as they evolve.

Next, we associated the temporal dynamics of healthcare unit occupancy alongside the laboratory-based surveillance indicators (Figure 2B-C). Notably, the increasing occupancy levels during the first and second waves tend to anticipate the peaks of respiratory pathogen outbreak progression by up to five weeks. Although the third wave demonstrated a reduced lead time for anticipating the pathogen surge, a substantial real-time increase in occupancy levels was observed following the increase in laboratory-confirmed cases. Even in scenarios where early anticipation is limited, such occupancy-based signals remain valuable for situational awareness and can support timely decision-making by healthcare managers. Also, this reduced anticipation window is partially explained by the decline in DENV circulation during the second semester of 2024, followed by a sudden rise in SARS-CoV-2 cases in August 2024 (Figure 2B-C). The transition between these two epidemiological phases created a pronounced valley in overall laboratory-based indicators around June and July.

### High occupancy averages indicate elevated laboratory-confirmed infections

To quantify temporal similarity between healthcare occupancy and confirmed pathogen-specific epidemiological events, we applied DTW on scaled frequency over all 66 weeks of data. Lower DTW distances indicate greater temporal similarity after allowing for local shifts in time, making it a suitable metric for comparing epidemic signals with potential lags or phase differences. SARS-CoV-2 and RV test positivity rate indicators showed the strongest temporal concordance with occupancy patterns, as reflected in their lower warping distances with 0.89 and 0.97, respectively. In contrast, DENV-related series showed higher distances, indicating phase shifts or misalignment relative to occupancy surges. Such distance was primarily driven by a single pronounced peak in dengue virus activity during the second half of the analyzed period. Because DTW was calculated over the full timeline, this abrupt and concentrated nature of the DENV wave reduced the overall similarity with the occupancy trends.

As the observed alignments from the DTW analysis demonstrated a lag structure, particularly the consistent 5-week offsets between the peaks (Figure 2B-C), we next evaluated whether changes in occupancy could statistically inform subsequent trends in pathogen-specific signals. To capture time-lagged influences across distinct epidemic dynamics, we opted to segment the data into three partially overlapping lag-aware periods: a first period from July to December 2023, a second from October 2023 to June 2024, and a third period from June 2023 to October 2024. The overlapping lag-aware periods account for the temporal lags specific to each pathogen while enabling clearer attribution of occupancy changes to individual epidemic signals. Directional dependencies between time series were further assessed using the Granger causality test applied to differenced and detrended data. Significant associations (p<0.05) were identified across all three shifted periods (Table 1). In the first lag-aware period (July–December 2023), we detected significant associations for all three pathogen groups, with RV-related indicators contributing the highest number of causal relationships (n=5), followed by DENV (n=2) and SARS-CoV-2 (n=2). There was a mean lag of 3 weeks across these associations, supporting the findings from the DTW analysis. During the second lag-aware period (October 2023–June 2024), DENV emerged as the dominant driver of occupancy variation, with five highly significant associations (p<0.01) and lags ranging from one to five weeks. No significant associations were observed for RV or SARS-CoV-2 during this period, suggesting that dengue activity alone accounted for occupancy increases during this phase. In the third lag-aware period (June–October 2024), signal strength was lower overall, but short-term directional associations (lag=1) were identified for both SARS-CoV-2 and RV indicators with marginal significance (p=0.03 and p=0.04, respectively). These wave-specific results reinforce the notion that healthcare occupancy responds to different pathogens and further demonstrate its potential as a lag-sensitive indicator of epidemic burden.

### Early warning of epidemic burden using occupancy-derived volatility metrics

On the basis of this temporal lag structure, we applied a 42-day (6-week) moving average to healthcare occupancy data to calculate the z-scores (Figure 4A). This window was selected based on the consistent 5-week lead time between occupancy data and pathogen-specific signals, with an additional week added to enhance sensitivity and minimize the influence of short-term noise. Weekly deviations (standard deviations, SD) from the moving average were defined as the indicator of system pressure. During the initial phase of each occupancy wave, the volatility signals intensify as weekly values rise above the moving average. This signal converges at the peak of the wave, where short-term and long-term trends align. The opposite occurs in the later stages of the detected events, where weekly occupancy values begin to decline while the moving average remains elevated, resulting in a signal indicative of decreasing healthcare system usage. Three distinct anomalies were evident in the deviations from the mean, beginning in August 2023, December 2023, and August 2024 (Figure 4A).

Although aggregated trends across all 17 units provide an overview of occupancy patterns in the analyzed region, the same volatility index can be disaggregated to understand unit-specific dynamics. When comparing the initial (August 26, 2023 - Figure 4B-D) against the later stages (November 04, 2023 - Figure 4E-G) of an outbreak, we identified distinct spatial and temporal in-unit occupancies. At the earlier time point, all units showed positive deviations from the moving average, including 13 with high-intensity alerts (red) and four with moderate deviations (yellow), with most facilities operating near or above 50% occupancy. By the later time point, 10 units exhibited occupancy declines (green bars), while the remaining 7 continued to show signs of strain. Importantly, 16 of the 17 units recorded occupancy rates below 50%, suggesting a system-wide reduction in demand.

Localized areas of persistent pressure were identified using the Local Indicators of Spatial Association (LISA) test, which identified clusters of units with similar occupancy patterns. Four units (8, 9, 10, and 15) revealed a significant hotspot (p-value=0.03) during the later phase of the outbreak (Figure 4G). Mapping these hotspots exposed regions with abnormally high occupancy during both non-outbreak (Supp. Figure 1A) and outbreak periods (Supp. Figure 1B, Supplementary Data 2). These findings demonstrate that, even as overall demand decreases, spatial heterogeneity in healthcare burden persists and highlight the importance of unit-level monitoring for targeted public health interventions.

## Discussion

While many disease forecasting models aim to predict pathogen-specific trends, each disease follows its own transmission dynamics, often limiting the generalizability of such approaches. However, from a surveillance management perspective, the priority is to identify anomalous activity early enough to allocate resources and mitigate the healthcare systemic pressure. Our study with sixty weeks of data during 2023-2024 proposes an innovative approach to track real-time healthcare occupancy and highlight periods of health system pressure. We demonstrated the use of occupancy data as a complementary system that harnesses alternative data streams to enhance the speed, sensitivity, and scalability of epidemic surveillance. Regardless of the disease etiology, the occupancy signal preceded the peak of cases of pathogens of major public health importance by up to five weeks, as indicated by the first occupancy wave. This temporal relationship was observed consistently across multiple epidemic waves and for distinct pathogen infections, reinforcing the idea of using occupancy trends for epidemiological purposes.

Although infections caused by viral pathogens tend to follow well-defined seasonal patterns, multiple factors can interfere with transmission dynamics and either advance or delay expected epidemiological events, such as climate change ^16^ and human behavioral shifts like lockdowns ^17^. For this reason, it is necessary to use a method sensitive enough to capture nuanced variations in health care systems. In parallel, there is an increasing demand from the Ministry of Health (MoH) to support efficient early warning systems to enhance evidence-informed epidemic management ^18^. Our work offers a scalable approach that indicates outbreak activity as a complement to the existing surveillance methods. During public health emergencies, this occupancy-based method can be used as an operational trigger for further epidemiological investigations to allocate resources for pathogen identification in a timely manner. This capability supports the development of an adaptive surveillance system that can detect emerging threats faster and aligns with the Brazilian MoH’s goal of improving the responsiveness of Brazil’s public health surveillance infrastructure.

The temporal associations observed between occupancy data and laboratory-based indicators offer strong support for the use of healthcare facility activity as a leading signal of epidemic burden. Importantly, the occupancy signal demonstrated a consistent ability to reflect underlying infection dynamics, whether through anticipation or real-time alignment, as seen in the third wave. This underscores the broader utility of occupancy trends not only for forecasting but also for near real-time situational awareness, particularly when traditional surveillance systems may lag or underperform. The sensitivity of this signal to both arboviral and respiratory pathogens, regardless of their transmission mechanisms or seasonality, underscores its integrative value for public health surveillance. Moreover, the detection of statistically significant lagged associations across multiple periods through Granger causality analysis supports the robustness and reproducibility of this approach in diverse epidemic contexts.

Due to its sensitivity to healthcare system burden regardless of disease etiology, the occupancy signal anticipated a 5-week lead between the first peak of occupancy and the peak of the second SARS-CoV-2 wave (Figure 2B). However, during the decline of this SARS-CoV-2 wave, occupancy levels did not decrease in parallel, likely due to the simultaneous increase in dengue virus (DENV) infections. Between 2023 and 2024, a total of 3.79 million dengue cases were reported in Brazil, with 52% occurring in the Southeast region ^19^. The state of São Paulo, the most densely populated area in the country, accounted for a significant share of this burden with 2,745 cases per 100,000 inhabitants. This represented the highest dengue burden during the period analyzed. Notably, the occupancy levels remained sensitive throughout, capturing the distinct dynamics of both SARS-CoV-2 and DENV waves despite their differing seasonality, further underscoring its value as a potential integrative and timely indicator of epidemic pressure.

The integration of a volatility-based metric with spatial association analysis provides an additional layer of resolution for identifying anomalous patterns in healthcare system usage. By capturing deviations from expected occupancy levels at both regional and unit-specific scales, this method adapts dynamically to reveal localized system pressures that may signal emerging outbreaks before they escalate to broader system-wide impact. Notably, occupancy surges in just a few units within a single municipality can already serve as actionable warning signals for local health administrators. These early alerts offer not only operational value but also strategic insights into structural disparities in healthcare access. Persistent high-occupancy hotspots revealed areas of concentrated demand and potential gaps in the equitable distribution of healthcare services. Recognizing these spatial inequities is crucial for guiding the reallocation of resources, including the redistribution of human resources and medical supplies to the most affected units. In this way, the volatility index, combined with spatial analysis, can support health system equity goals by identifying zones where population needs are highest and response capacity may be insufficient. This strengthens the capacity for targeted interventions, enabling managers to balance efficiency with fairness in epidemic preparedness and response.

Despite the strengths of this approach, it has limitations. Notably, the Google Maps™ occupancy data lacks full detailed methodological transparency and may be subject to representativeness bias depending on the type of health care unit, user demographics, and regional coverage. Therefore, while this method offers valuable insights into the health care system burden, it is not intended to work in isolation. Its full potential is realized when integrated with other surveillance approaches such as syndromic, laboratory, and genomic surveillance. These classical epidemiological approaches provide pathogen-specific and population-level contextual information. Together, these complementary tools can enhance the resolution of health surveillance and public health policies.

### Conclusions

This study presented an innovative, low-cost, and easy-to-implement method with the capability to detect epidemiological events up to five weeks in advance compared to laboratory test data. Our approach provides valuable insights that can support public health managers in making data-driven decisions and implementing effective control measures. The geospatial visualization and comparative analysis between outbreak and non-outbreak periods emphasize the importance of continuous surveillance and efficient health service management to prevent system overburden and ensure quality care for the population. Future improvements should focus on expanding the epidemiological data pool and incorporating seasonal variables for an even more robust approach. These findings underscore the relevance of real-time data monitoring as a critical component of surveillance systems capable of timely epidemic response.

## Methods

### Data source and selection criteria

Real-time occupancy percentage values were obtained from Google Maps™ from 17 emergency care units hourly from seven cities in the São Paulo metropolitan area between July 15, 2023, and October 12, 2024. During the collection process, we ensured that no personal information was obtained and all data were aggregated to ensure safety compliance. These units were randomly selected to represent different neighborhoods in the São Paulo area, which also serves as a primary hub for the state. The use of metropolitan area data as representative of broader regional and state patterns is supported by several studies that have demonstrated significant associations between metropolitan and state-level epidemiological indicators ^20–22^. Five epidemiological weeks (2023-08-19, 2023-11-25, 2023-12-02, 2023-12-09, 2023-12-30) were excluded due to consecutive days with missing data. In total, 66 weeks were available for subsequent analysis. For weeks with only a single missing hourly record, numerical data interpolation (method=’linear’) was applied. These non-consecutive missing data points typically resulted from interruptions in internet connectivity.

Diagnostic test results for SARS-CoV-2, influenza virus (A and B), respiratory syncytial virus, and dengue virus were obtained from different sources. First, anonymized diagnostic test data were obtained from seven private laboratories that compose a prospective pathogen monitoring initiative coordinated by the Instituto Todos pela Saúde (ITpS) - All for Health Institute ^23^. These diagnostic data are mainly derived from symptomatic individuals seeking assistance in private healthcare facilities in São Paulo state. In this case, randomly selected individuals, including symptomatic and asymptomatic individuals, were tested. Second, test results were obtained from the government epidemiological surveillance information system SIVEP-SRAG from Open Datasus, which monitors cases of Severe Acute Respiratory Infections (SARI). SIVEP-SRAG is a compulsory notification repository for COVID-19 and other respiratory diseases from both public and private healthcare systems in Brazil. Third, DENV case data were sourced from InfoDengue ^24^, a surveillance platform that issues arbovirus alerts. In this study, we included only confirmed cases with pathogen detection, represented by the number of positive results from antigen or RT-qPCR tests from the three data sources. All datasets used in this study (ITpS, SIVEP-SRAG, and InfoDengue) are from public repositories that do not provide sensitive patient data; therefore, analyses were based on them, follow open data principles, and do not require ethics committee approval in Brazil. Real-time occupancy and laboratory data were consolidated for downstream epidemiological analyses (Supplementary Data 3)

### Time series similarity assessment

Hourly data from each healthcare facility were first aggregated into daily averages. Subsequently, the weekly occupancy values were calculated as the average of the daily values of each epidemiological week. We separated the laboratory-based surveillance data by source and three main groups: (i) SARS-CoV-2, (ii) DENV, and (iii) a respiratory panel of important pathogens for Brazilian public health, including SARS-CoV-2, respiratory syncytial virus (RSV), Influenza A and B. These diagnostic test results were consolidated weekly. For disease case counts from SIVEP-SRAG and InfoDengue public repositories, we used the number of positive tests. For infection rates, we calculated the positivity rate by dividing the number of positive tests by the total number of tests and multiplying by 100 to obtain the percentage.

Dynamic time warping from the dtaidistance library [v.2.3.12] ^25^ was applied to assess the temporal alignment between the occupancy data and the laboratory-based surveillance data. By default, DTW returns a distance of zero when comparing a time series with itself, and increases as temporal misalignment or shape differences grow. This makes it suitable for capturing non-linear shifts and local distortions in timing between epidemic indicators. To ensure comparability of temporal patterns rather than absolute magnitudes, each time series was normalized independently by linearly rescaling its minimum and maximum values to a 0-1 range using scikit-learn ^26^. Given the multiple pathogen time series analyzed, we adjusted the occupancy wave periods according to the observed temporal lags in pathogen circulation to ensure evaluation of the complete trend period. The partial overlaps introduced by this adjustment were identified using DTW, which aligns temporal patterns across datasets and highlights phase shifts between signals.

To evaluate whether temporal changes could consistently explain the relationship between emergency care unit occupancy and seasonal variations in pathogen circulation, we applied the Granger causality test. The Augmented Dickey-Fuller (ADF) test was used to evaluate stationarity, and first-order differencing was applied when necessary. We analyzed three distinct temporal waves defined based on epidemiological weeks. For each wave, we tested the causality between occupancy percentage changes and pathogen-specific indicators using the statsmodels [v.0.14.4] ^27^ implementation of the Granger causality test, considering lags tested up to five weeks due to data availability constraints on each wave. All statistical estimates, such as lag-specific and p-values, were exported for further interpretation.

### Z-based epidemic volatility index

Next, to monitor fluctuations in emergency care unit occupancy, we adapted the epidemic volatility index ^28^, an early warning algorithm designed to detect emerging epidemic waves by assessing relative changes in the standard deviation of case counts over time. In our adaptation, hourly occupancy data were aggregated into daily averages. For each facility, we computed a 42-day moving average and standard deviation, using a window length set one week longer than the maximum lag tested in the Granger causality analyses to ensure short-term trends were represented. Z-scores or standard deviations from the mean were calculated by subtracting the moving average from the daily occupancy percentage and dividing by the corresponding standard deviation. These daily z-scores were then aggregated into weekly summaries aligned with epidemiological weeks. To classify the intensity of occupancy fluctuations, we applied predefined thresholds to the weekly z-scores. Values below 0 were labeled indicating low volatility (green), values between 0 and 0.65 as moderate volatility (yellow), and values above 0.65 (high volatility). We selected this threshold for high volatility as it represents a moderate deviation - approximately two-thirds of a standard deviation-from the moving average over a 42-day (six weeks) window. Under a normal distribution, about 74% of observations fall below 0.65 standard deviations, making this threshold sensitive enough to detect atypical increases in occupancy while limiting false positives from routine variability. This choice also accounts for the limited range of percentage data, which reduces variability compared to absolute case counts.

### Geospatial statistics information

Next, to evaluate spatial relationships between healthcare units, we computed pairwise distances using Haversine’s algorithm to generate a geographic distance matrix (here in kilometers) across each pair of units (Supp. Figure 2). This algorithm calculates circle distances based on latitude and longitude coordinates while accounting for the Earth’s curvature. To identify localized patterns of weekly occupancy, we applied the Local Indicators of Spatial Association (LISA) test to average occupancy rates vs geographical distance. The LISA method detects spatial clusters of similar or dissimilar values by comparing each unit’s occupancy value with those of its nearest neighbors. For this, we used a spatial weights matrix based on the nearest neighbors (k=4) with significant units assigned witha p-value<0.05. Geospatial maps and metrics were visualized using GeoPandas [v.1.0.1] and Matplotlib [v.3.10.0].

## Data Availability

All data produced in the present work are contained in the manuscript

https://github.com/InstitutoTodosPelaSaude/paper_occupancy_detecta

## Acknowledgements

We acknowledge Google Maps™ for providing aggregate occupancy data essential to this study. We are grateful to the Brazilian Ministry of Health for public access to SIVEP-SRAG and InfoDengue surveillance platforms. The findings and conclusions are solely those of the authors and do not necessarily reflect the official position of the funding institution.

## Authorship information

J.D.A., J.C.S.S., and V.S.S. contributed to the conceptualization and methodology of the study. E.R.S., J.D.A., and J.C.S.S. were responsible for software development and data curation. Validation was performed by J.D.A., J.C.S.S., M.A.S.B., E.C.S., H.I.N., C.S.L., J.R.R.P., G.O.P. and V.S.S. Formal analysis and investigation were carried out by J.D.A., J.C.S.S. and V.S.S. A.F.B. and M.S. provided resources. Data visualization was conducted by I.N.S., J.D.A., J.C.S.S., and A.F.B. The original draft of the manuscript was written by J.D.A., J.C.S.S., and V.S.S., and review and editing were performed by J.D.A., J.C.S.S., M.A.S.B., C.S.L., J.R.R.P., A.F.B,. and V.S.S. Supervision was provided by J.D.A., J.C.S.S., E.C.S., H.I.N., G.O.P., J.K., M.S., A.F.B,. and V.S. Project administration was conducted by J.D.A., J.C.S.S., G.O.P., J.K., M.S., and V.S. Funding acquisition was carried out by J.D.A., J.C.S.S., G.O.P., J.K., M.S. and V.S.S.

## Data and code availability

The consolidated datasets and the complete Python codes to perform the analyses of this work are available at https://github.com/InstitutoTodosPelaSaude/paper_occupancy_detecta.

## Declaration of Interests

The authors declare no competing interests.

---

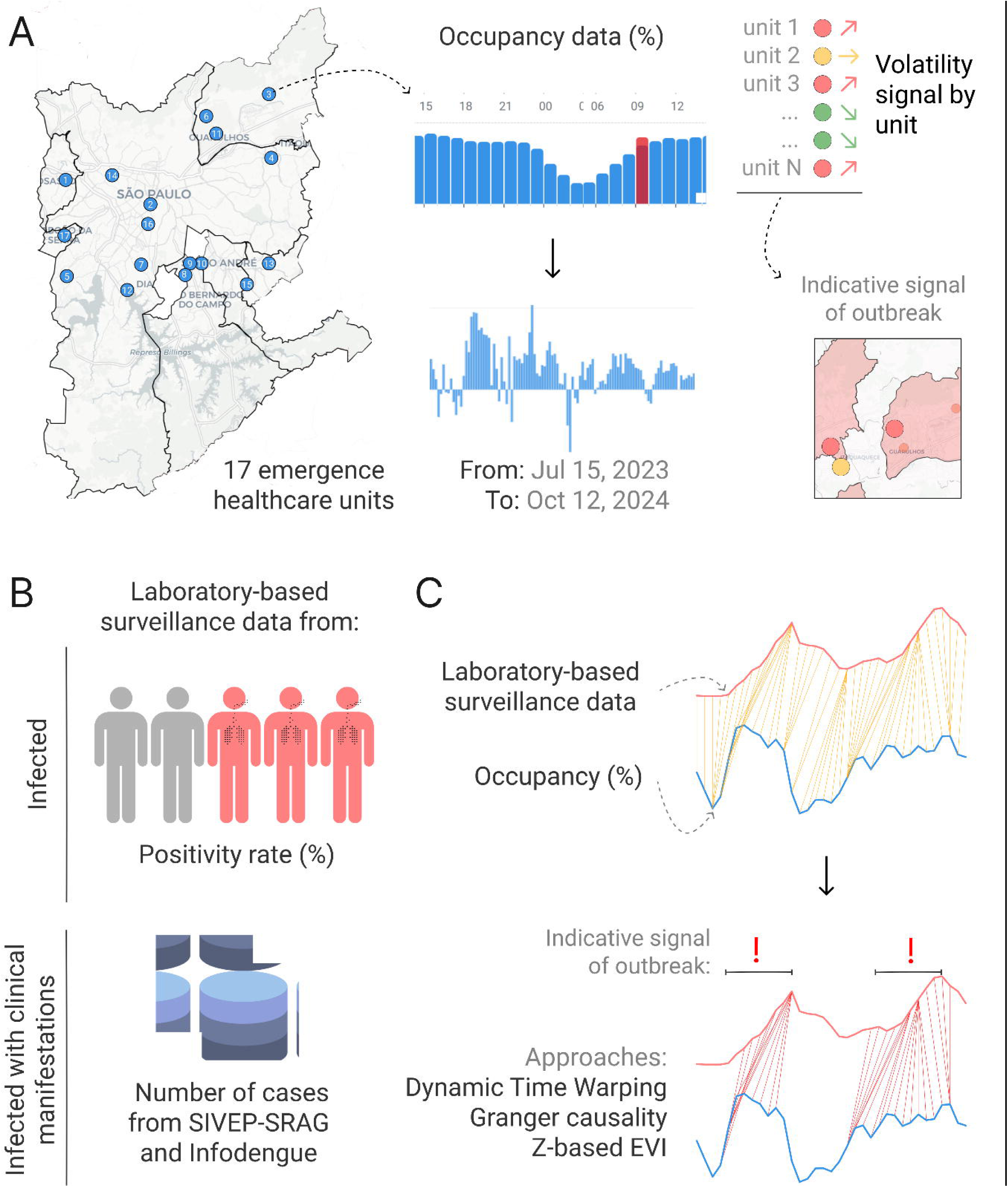

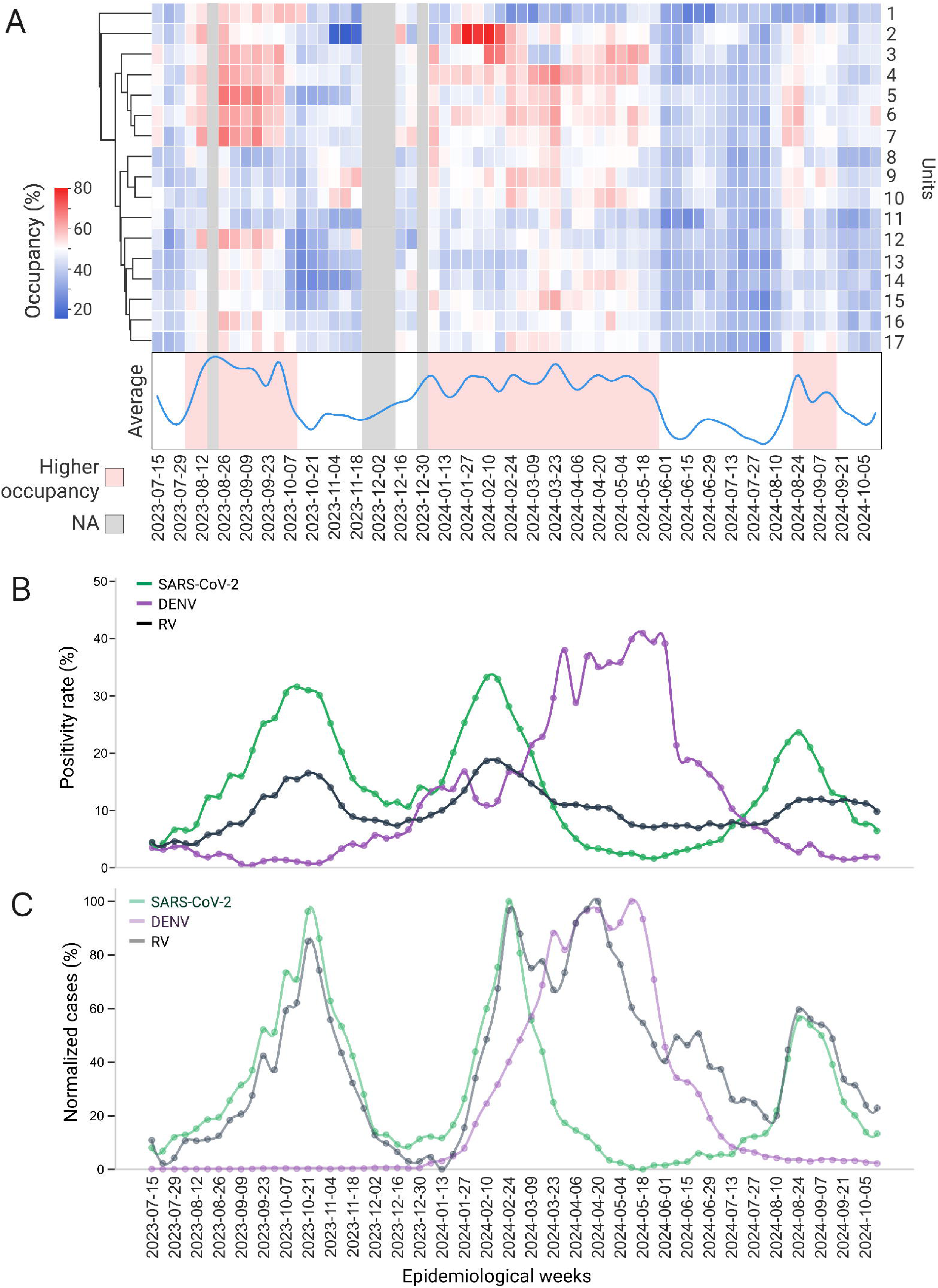

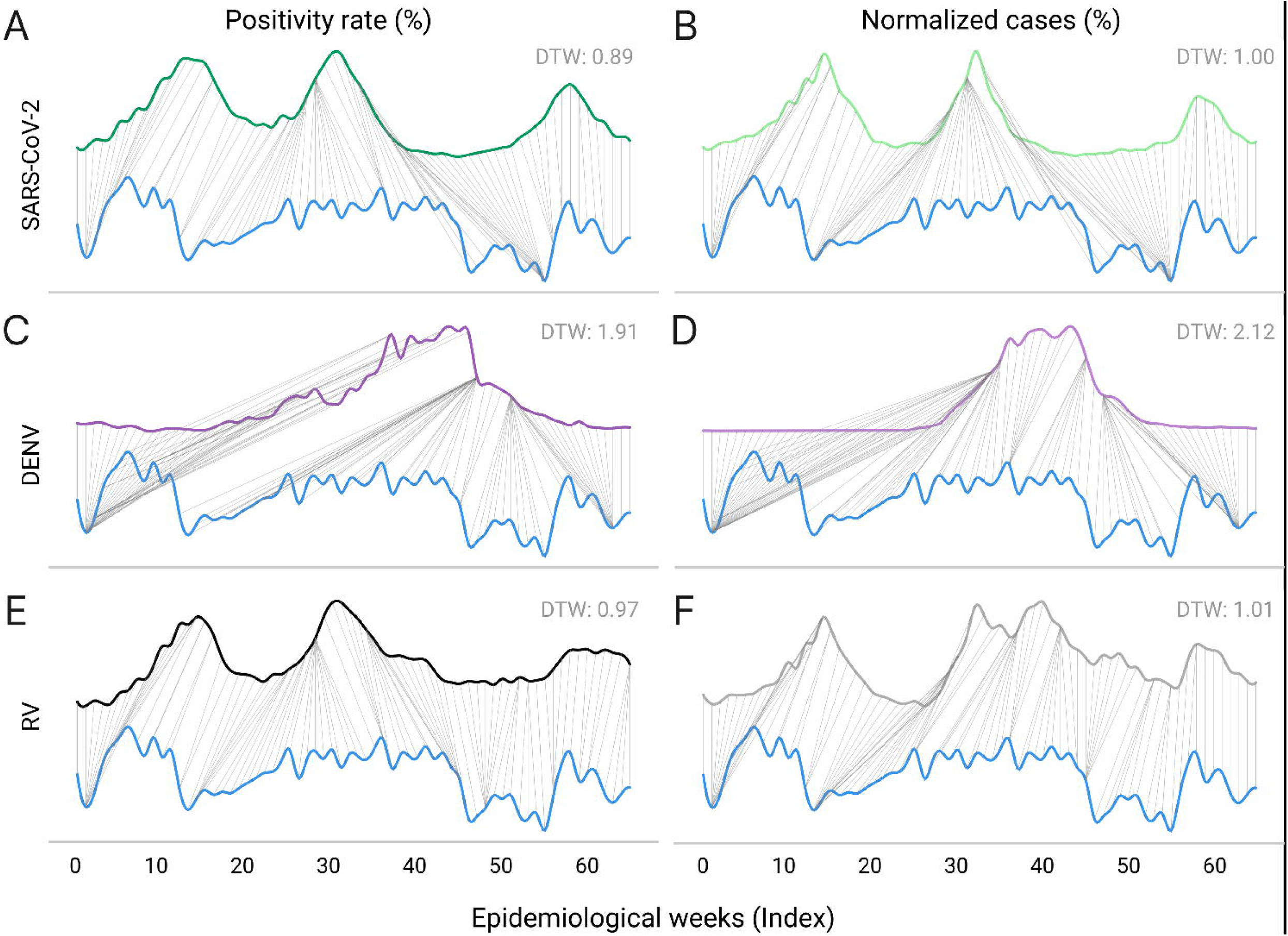

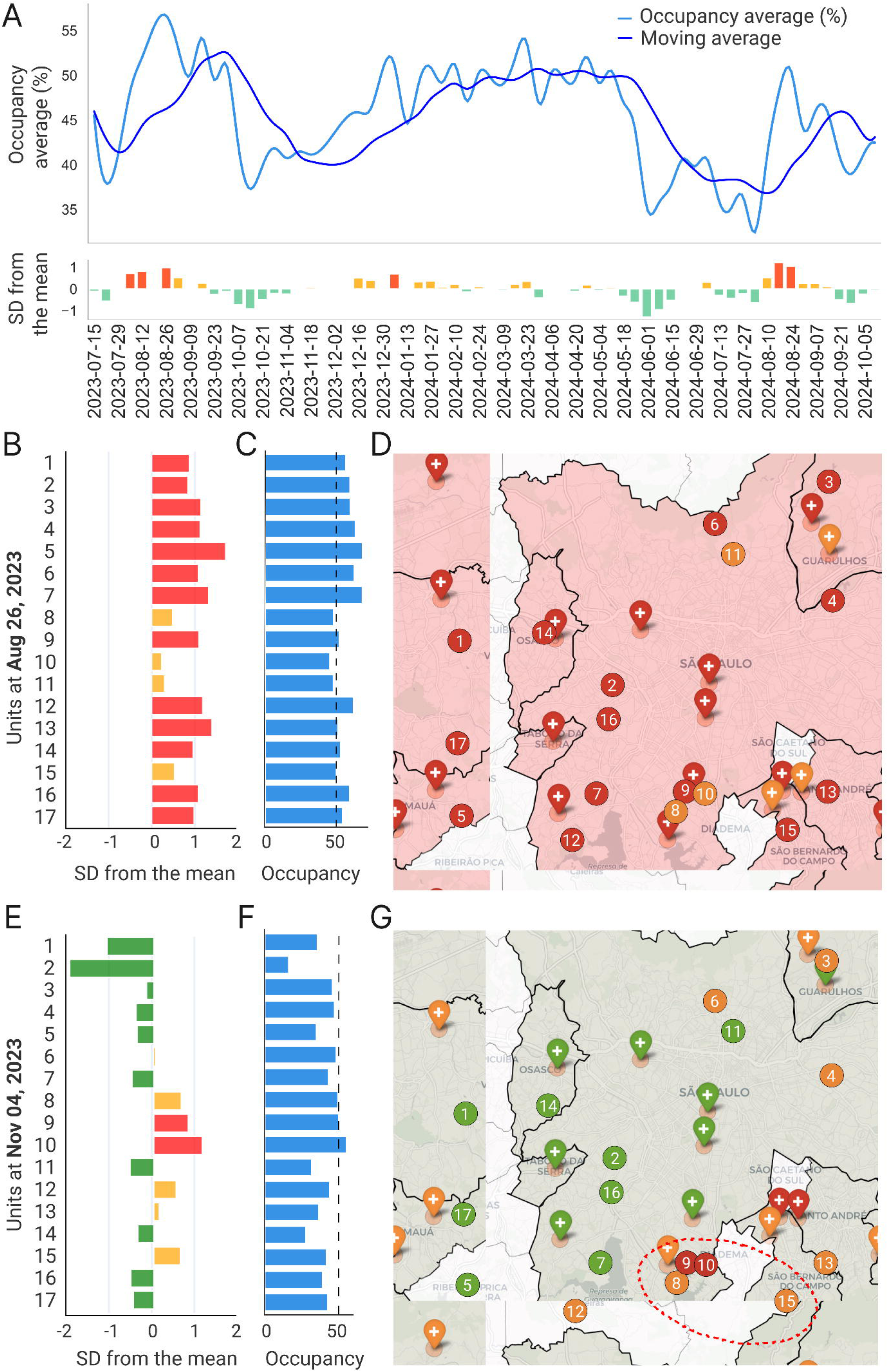

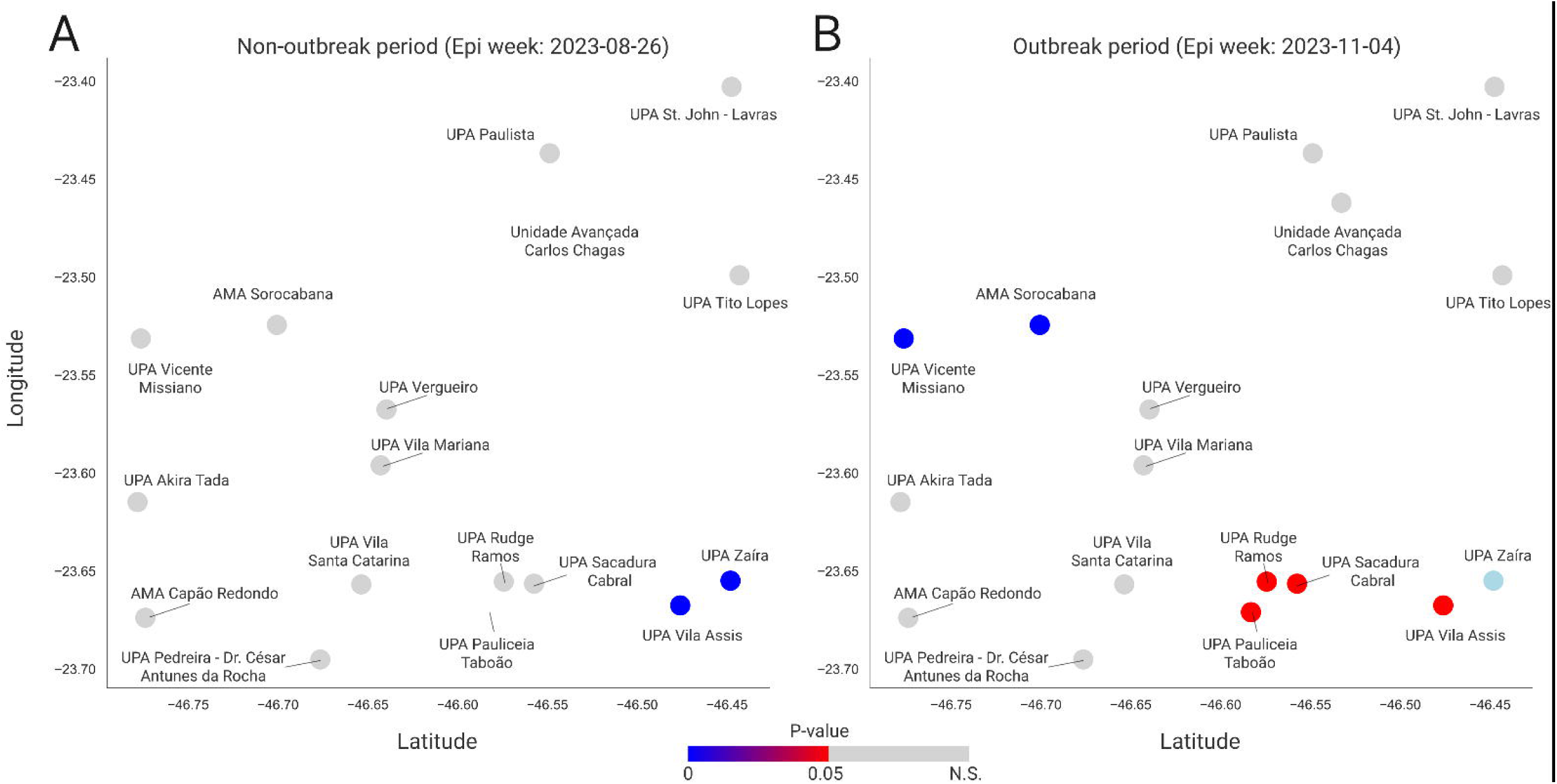

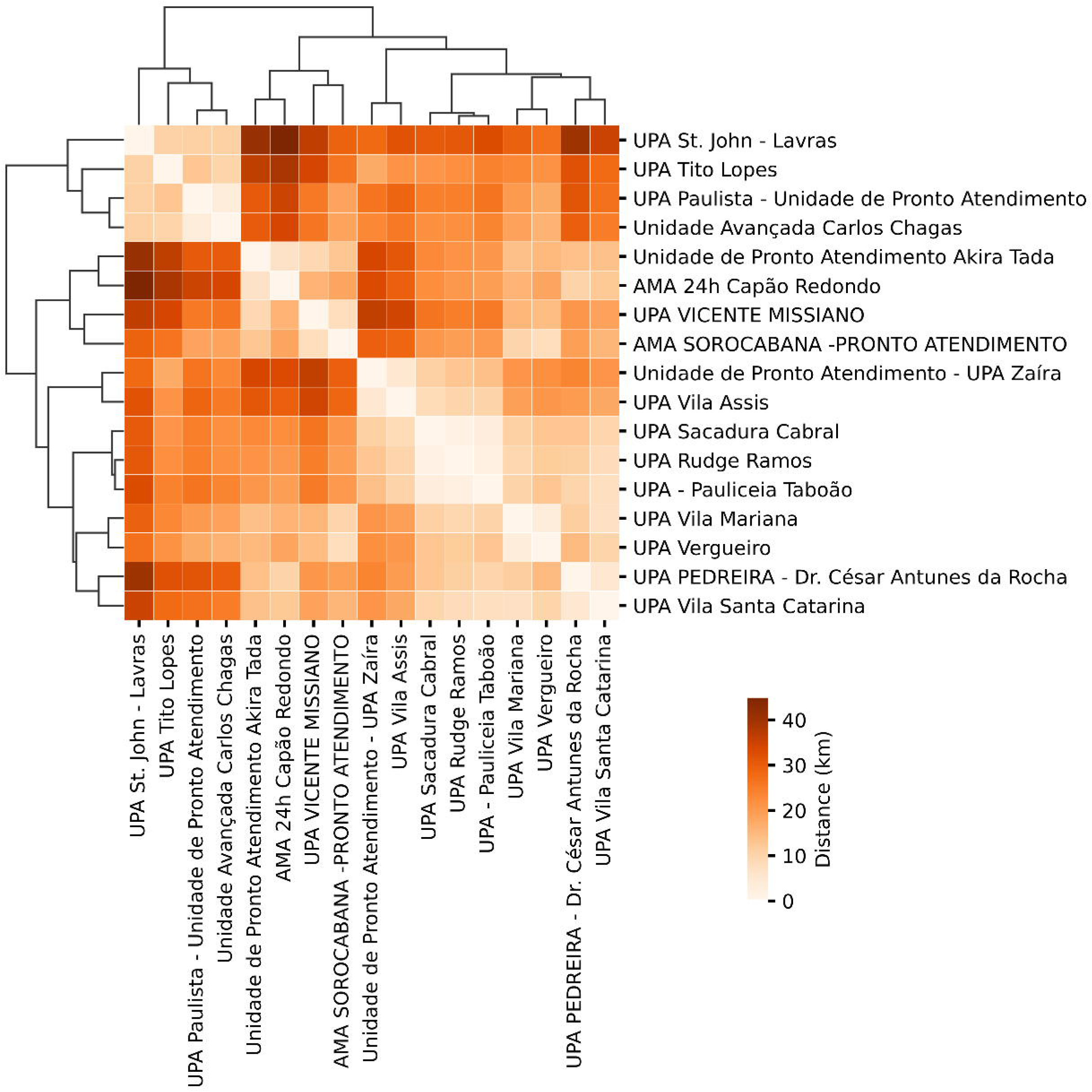

## Notes

### Competing Interest Statement

The authors have declared no competing interest.

### Funding Statement

This study was funded by ITpS.
VSS has a fellowship from CNPq-PQ 2024.

